# Myocardial T1 and T2 Mapping at 5.0 T: Feasibility and Comparison with 3.0-T MRI

**DOI:** 10.1101/2025.08.03.25332892

**Authors:** Qinfang Miao, Hongzhang Huang, Zhenfeng Lyu, Wenjian Liu, Tianyi Wu, Peng Hu, Haikun Qi

## Abstract

**Background:** Myocardial T1 and T2 mapping provide a non-invasive quantitative assessment of cardiac tissue. While established at 1.5-T and 3.0-T MRI, the mapping performance and reference values at 5.0-T MRI remain to be investigated. This study aims to evaluate the feasibility and establish reference values for cardiac T1 and T2 mapping at 5.0-T MRI in healthy subjects.

**Methods:** In this prospective study, 50 healthy volunteers underwent cardiac MRI at both 3.0-T and 5.0-T MRI systems from February to April 2025. Mapping protocols included MOdified Look Locker Inversion recovery (MOLLI, T1), T2-prepared gradient-echo (T2-prep-GRE, T2), and a simultaneous multi-parametric mapping technique (Multimap, T1 and T2). At 3.0-T MRI, both balanced steady-state free precession (bSSFP) and fast low-angle shot (FLASH) readouts were employed, while only the FLASH readout was used at 5.0-T MRI. Intra-observer, inter-observer, and scan-rescan reproducibility were assessed. Statistical analyses included coefficient of variation (CoV), intraclass correlation coefficient (ICC), and Bland-Altman analysis.

**Results:** All techniques at 5.0-T MRI yielded high-quality, artifact-free images. Native T1 values were significantly higher at 5.0 T than at 3.0-T MRI (MOLLI: 1452.9 ± 33.2 ms vs. 1305.9 ± 38.3 ms, *P* < 0.0001), while T2 values were significantly lower (T2-prep-GRE: 37.53 ± 1.74 ms vs. 43.97 ± 2.95 ms, *P* < 0.0001). Scan-rescan reproducibility at 5.0 T (ICC: 0.87–0.92; CoV: 2.41%–3.25%) was comparable to 3.0-T MRI. The measurement precision of 5.0 T was higher than 3.0-T MRI with FLASH readout and slightly inferior to bSSFP-based techniques. Multimap achieved efficient, simultaneous T1 and T2 quantification with acceptable reproducibility at 5.0-T MRI.

**Conclusion:** Myocardial T1 and T2 mapping at 5.0-T MRI are reliable and reproducible in healthy individuals, offering reference values for normal myocardium at this field strength. The good measurement reproducibility and precision of 5.0-T cardiac mapping support its clinical potential for myocardial tissue characterization.

## Background

Quantitative cardiac magnetic resonance (CMR) parametric imaging provides a non-invasive tool for assessing myocardial tissue and diagnosing cardiovascular diseases [1–3]. T1 and T2 mapping enable quantitative and objective evaluation of subtle pathological changes in the myocardium by directly measuring myocardial T1 and T2 relaxation times. Native T1 mapping can reflect changes related to inflammation, fibrosis, and infiltrative diseases [4–6]. T2 mapping is sensitive to myocardial edema and associated inflammatory response [7,8]. While conventional techniques quantify a single parameter in a breath-hold acquisition [9,10], recent technical developments enable the measurement of multiple parameters simultaneously in a single scan, improving the acquisition efficiency. Especially, the multi-parametric mapping techniques (Multimap) based on single-shot Cartesian acquisition and dictionary matching enable simultaneous quantification of multiple parameters in a short acquisition without the need for complex post-processing, and thus hold great promise for wide clinical adoption [11–13].

Cardiac T1 and T2 mapping have been well-established on 1.5-T or 3.0-T MR using electrocardiogram-triggered single-shot acquisition with balanced steady-state free precession (bSSFP) readout due to its high signal-to-noise ratio (SNR) efficiency. With the introduction of a whole-body 5.0-T scanner, there is increasing interest in performing CMR at this ultrahigh field strength [14–18], where higher spatial resolution or SNR can be expected. However, the sensitivity of bSSFP to off-resonance effects leads to banding artifacts in the presence of non-uniform B0, which hinders its adoption at 5.0 T due to the increased B0 inhomogeneity. Fast low-angle shot acquisition (FLASH) is more robust to B0 inhomogeneity, albeit with lower SNR than bSSFP. The feasibility of adopting FLASH readout for cardiac parametric mapping at 5.0 T, and the resultant precision and reproducibility have not been systematically evaluated.

The aim of this study is to assess the feasibility of myocardial mapping at 5.0 T by comparing the performance of T1, T2, and Multimap mapping techniques between 3.0-T and 5.0-T MRI in healthy volunteers, and establish 5.0-T system myocardial T1 and T2 reference values.

## Methods

### Study Design and Participants

This prospective study adhered to the Declaration of Helsinki and was approved by our institutional ethics committee. From February to April 2025, 53 healthy volunteers aged 18-65 years without cardiovascular diseases or symptoms were consecutively recruited. Exclusion criteria were contraindications to MRI, known systemic diseases, hearing impairment, and voluntary withdrawal. All participants provided written informed consent and then underwent cine CMR screening to confirm normal cardiac chamber size, wall motion, ejection fraction, and ventricular mass before final inclusion. All the recruited subjects completed CMR scans on both 3.0-T and 5.0-T MR with an interval of 4-24 hours between scans. To evaluate the scan-rescan reproducibility of T1 and T2 mapping, the identical CMR protocol was repeated respectively on the 3.0-T and 5.0-T scanner within 72 hours after the initial scan.

### CMR Acquisition Protocol

All examinations were performed on a 5.0-T MR scanner (uMR Jupiter, United Imaging Healthcare, Shanghai, China) equipped with a 24-channel body coil and a 24-channel spine coil, and a 3.0-T MR scanner (uMR890, United Imaging Healthcare, Shanghai, China) equipped with a 12-channel body coil and a 24-channel spine coil.

The CMR protocol included Cine for cardiac function assessment and T1 and T2 mapping acquired at basal, middle, and apical short-axis positions of the left ventricle. The employed mapping techniques included widely adopted MOdified Look Locker Inversion recovery (MOLLI) 5-(3)-3 for T1 [9], T2-prepared Gradient Echo (T2-prep-GRE) for T2 [10], and Multimap for simultaneous quantification of T1 and T2 in a single acquisition of 10 heartbeats where inversion recovery pulse is applied at the first and fifth cardiac cycle and T2-prep module is performed at the last three cardiac cycles [11]. The FLASH readout was adopted for the 5.0-T mapping acquisitions, while both bSSFP and FLASH readouts were adopted for the mapping scans at 3.0 T. All mapping sequences utilized electrocardiogram triggering to acquire images at end-diastole under breath-hold. For fair comparison, all mapping sequences shared the same spatial imaging parameters across both field strength systems, including voxel size = 1.67 mm × 2.08 mm, field of view = 320 mm × 280 mm, slice thickness = 8 mm, and GRAPPA = 2. The parameter settings of the FLASH acquisition were the same between the 3.0-T and 5.0-T systems. Detailed imaging parameters of the quantitative CMR sequences are provided in Table 1.

**Table 1:**
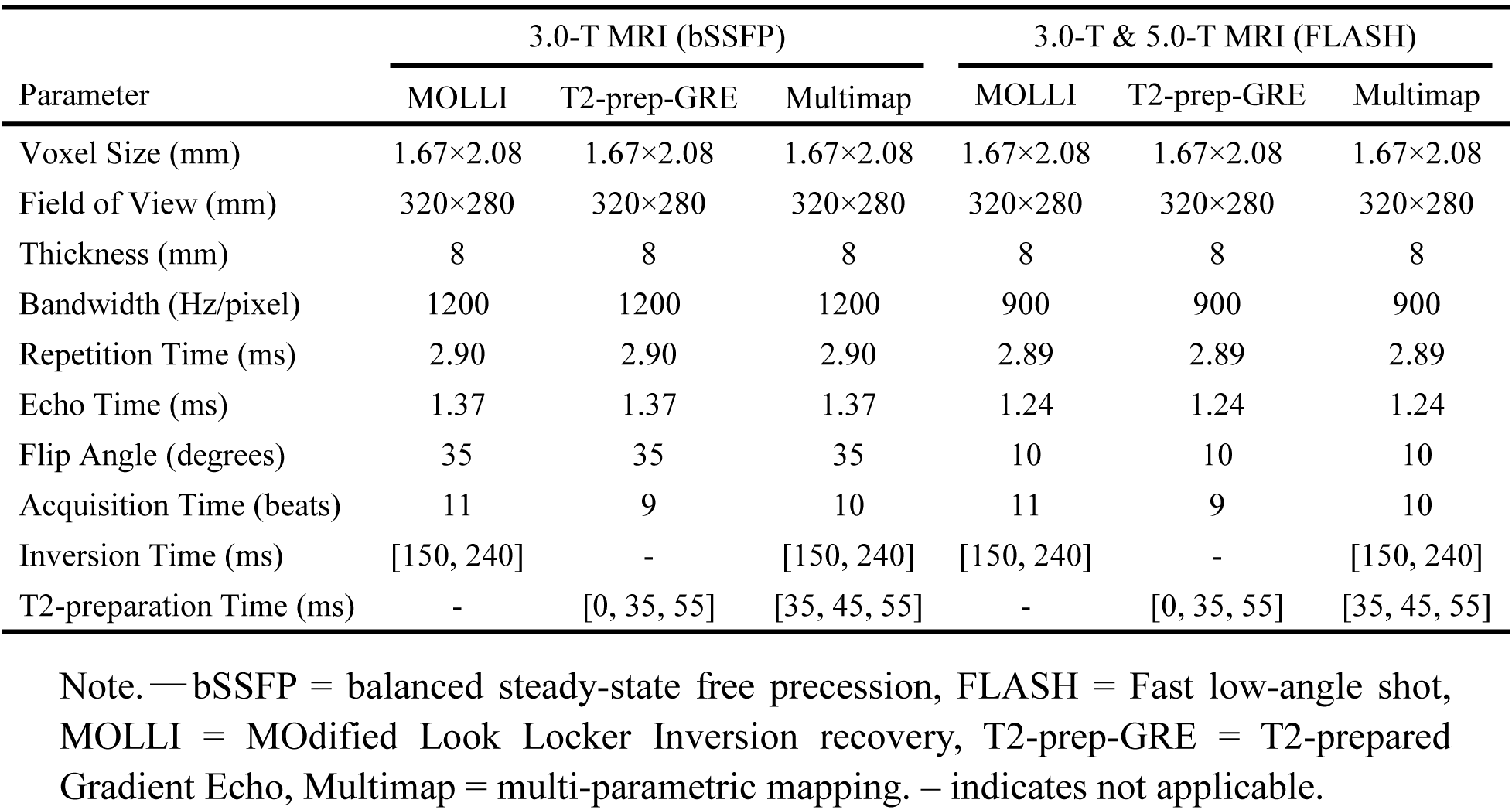
Imaging Parameters for the 3.0-T and 5.0-T Quantitative Cardiac MR Sequences.

| Parameter | 3.0-T MRI (bSSFP) |  |  | 3.0-T & 5.0-T MRI (FLASH) |  |  |
| --- | --- | --- | --- | --- | --- | --- |
|  | MOLLI | T2-prep-GRE | Multimap | MOLLI | T2-prep-GRE | Multimap |
| Voxel Size (mm) | 1.67×2.08 | 1.67×2.08 | 1.67×2.08 | 1.67×2.08 | 1.67×2.08 | 1.67×2.08 |
| Field of View (mm) | 320×280 | 320×280 | 320×280 | 320×280 | 320×280 | 320×280 |
| Thickness (mm) | 8 | 8 | 8 | 8 | 8 | 8 |
| Bandwidth (Hz/pixel) | 1200 | 1200 | 1200 | 900 | 900 | 900 |
| Repetition Time (ms) | 2.90 | 2.90 | 2.90 | 2.89 | 2.89 | 2.89 |
| Echo Time (ms) | 1.37 | 1.37 | 1.37 | 1.24 | 1.24 | 1.24 |
| Flip Angle (degrees) | 35 | 35 | 35 | 10 | 10 | 10 |
| Acquisition Time (beats) | 11 | 9 | 10 | 11 | 9 | 10 |
| Inversion Time (ms) | [150, 240] | - | [150, 240] | [150, 240] | - | [150, 240] |
| T2-preparation Time (ms) | - | [0, 35, 55] | [35, 45, 55] | - | [0, 35, 55] | [35, 45, 55] |

Note. —bSSFP = balanced steady-state free precession, FLASH = Fast low-angle shot, MOLLI = MODified Look Locker Inversion recovery, T2-prep-GRE = T2-prepared Gradient Echo, Multimap = multi-parametric mapping. – indicates not applicable.

### Image Processing

Cine images were analyzed using commercial CMR post-processing software (MASS, Medis, Leiden University Medical Centre, Leiden, The Netherlands) for measuring ejection fraction, ventricular mass and body surface area-indexed myocardial mass. The automatically generated myocardial contours were reviewed and manually corrected to ensure the accuracy of cardiac function measurements.

The T1 and T2 mapping acquisitions were processed and analyzed offline using MATLAB (R2022a, MathWorks, Inc., Natick, Massachusetts, USA). Firstly, non-rigid registration (17) was applied to the multi-contrast images to correct potential inter-contrast misalignments. Secondly, for quantitative mapping, bSSFP MOLLI and T2-prep-GRE with both bSSFP and FLASH readouts respectively went through standard three-parameter and two-parameter model fitting. For FLASH MOLLI and Multimap where the conventional analytical parameter model is not applicable, dictionary matching was performed to derive T1/T2 values. The subject-specific dictionary was generated by simulating the signal evolution during the acquisition based on the Bloch equation, incorporating the recorded R-R intervals (the time between consecutive R waves in the electrocardiogram) and trigger delay information. Besides T1 and T2, the B1+ spin history was modelled during dictionary simulation to improve the measurement accuracy with the following settings [11]: T1, [400:20:800, 805:5:1600, 1620:20:2200] ms; T2, [2:2:70, 75:5:120] ms; B1+, [0.4:0.04:1.2].

### Analysis of Quantitative Maps

The derived T1 and T2 maps were independently evaluated by two experienced CMR researchers (Q.M. and H.H.), who were blinded to participant information and the adopted magnetic field strength for imaging. The endocardial and epicardial borders were manually delineated on the T1 and T2 maps to define the myocardial region of interest, while avoiding interference from blood pools and surrounding fat. The mean and standard deviation (SD) were calculated for each short-axis slice to assess measurement accuracy and precision. The parameter distribution across the left ventricle was also assessed by performing segmental analysis according to the American Heart Association 16-segment model [21].

### Reproducibility Assessment

Intra-observer repeatability was assessed by having one observer (Q.M.) repeat the quantitative map analysis two weeks after the first analysis. Inter-observer reproducibility was evaluated by comparing the results of independent analyses of the same images by the two independent readers. Scan-rescan reproducibility was assessed by comparing data from repeated scans performed at the same field strength within 72 hours after the initial scan for all participants.

### Statistical Analysis

Statistical analysis and visualization were performed using Python (version 3.10, Python Software Foundation, Wilmington, Delaware, USA) and custom MATLAB scripts. Data normality was assessed using the Shapiro-Wilk test. Normally distributed continuous variables were presented as mean ± SD, while non-normally distributed data were presented as median and interquartile range. Categorical variables were expressed as frequencies (n, %). Group comparisons were performed using one-way analysis of variance with Bonferroni correction for multiple comparisons. Measurement precision was evaluated using the coefficient of variation (CoV, %). Reproducibility was quantified by calculating the intraclass correlation coefficient (ICC) with a two-way random-effects model, including the intra-observer repeatability, inter-observer reproducibility, and scan-rescan reproducibility. Systematic differences were assessed using Bland-Altman analysis, calculating mean differences (bias) and 95% limits of agreement (LOA). All statistical tests were two-sided, with significance set at *P* < 0.05.

## Results

### Characteristics of the Participants

Fig. 1A illustrates the study flowchart and inclusion criteria. Of the recruited 53 healthy volunteers, 50 participants (25 males, 25 females; mean age 44.0 ± 7.0 years) were ultimately included in this study, who successfully completed both 3.0-T and 5.0-T MR scans. The workflow of CMR examinations on the two scanners is visualized in Fig. 1B. The 3 subjects were excluded due to cardiac function abnormalities as detected by Cine CMR (n = 2), or voluntary withdrawal from the study (n = 1). Table 2 summarizes the participants’ demographic characteristics and cardiac structural and functional parameters. The age distribution was similar between male and female participants (males 44.4±7.0 years, females 43.6 ± 7.2 years, *P* = 0.68). There was also no significant difference in body mass index (females 23.5 ± 3.8 kg/m², males 24.5 ± 3.2 kg/m², *P* = 0.32), although the males had significantly larger body surface area than the females (1.92 ± 0.16 vs. 1.68 ± 0.15 m², *P* < 0.001). All participants had normal chamber size, normal wall motion, and ejection fraction (males 62.0 ± 5.3%, females 66.0 ± 4.8%, *P* = 0.007).

**Fig. 1.**
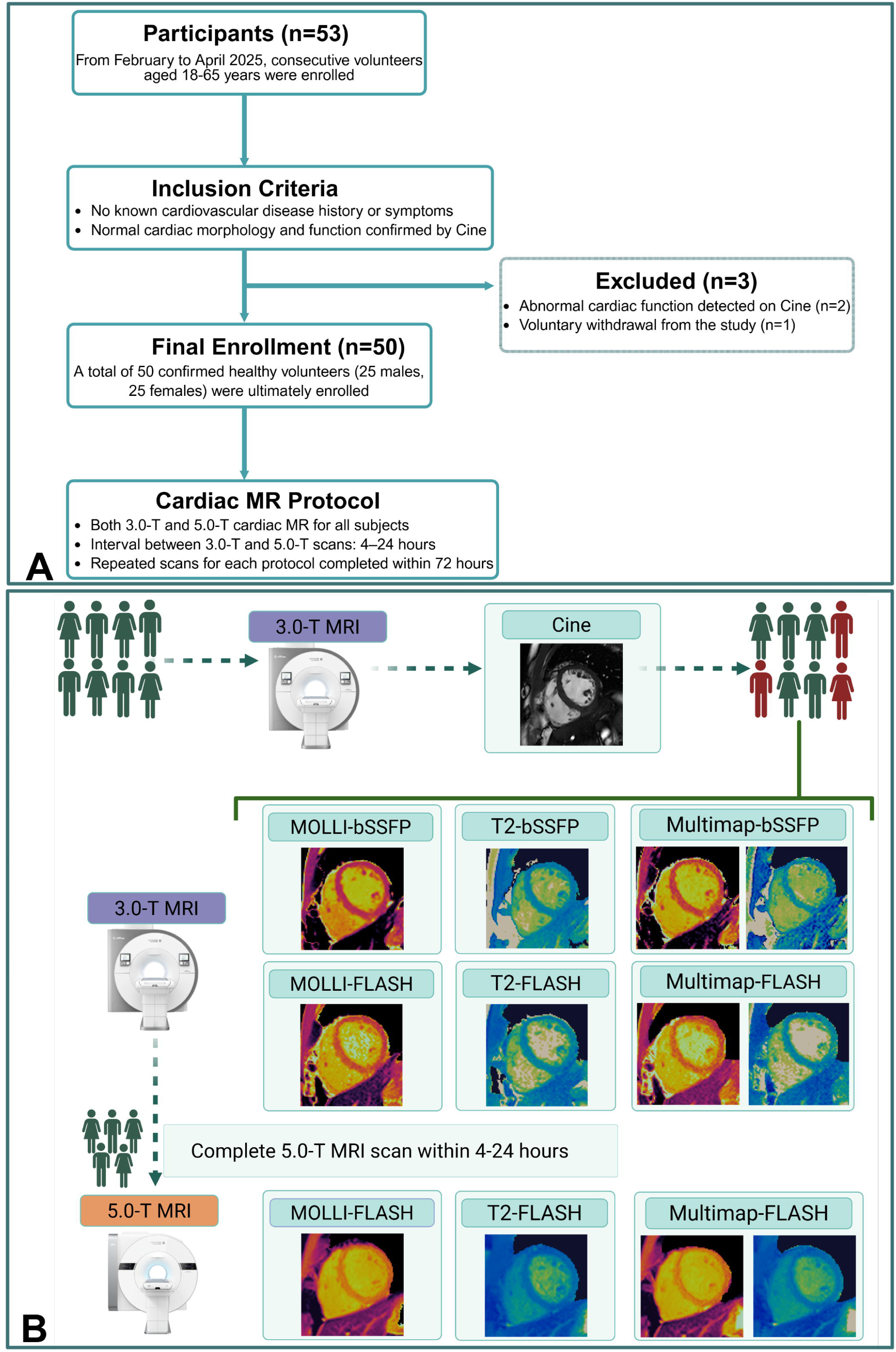
Study Design and Cardiac MR Protocol. **(A)** The study flowchart shows participant recruitment, inclusion criteria, and study protocol. Of the 53 initially recruited subjects, 3 were excluded due to abnormal cardiac function (n = 2) or voluntary withdrawal from the study (n = 1), resulting in 50 participants completing the entire protocol. **(B)** The imaging workflow illustrates the cardiac T1 and T2 mapping at both field strengths. At 3.0-T MRI, both bSSFP- and FLASH-based sequences were performed, including MOLLI, T2-prep-GRE, and Multimap. At 5.0-T MRI, only FLASH-based sequences were used due to magnetic field inhomogeneity and specific absorption rate limitations. MOLLI = MOdified Look Locker Inversion recovery, bSSFP = balanced steady-state free precession, Multimap = multi-parametric mapping, T2-prep-GRE = T2-prepared Gradient Echo, FLASH = Fast low-angle shot.

**Table 2:** Participant Characteristics.

| Characteristic | Female (*n* = 25) | Male (*n* = 25) | *P* Value |
| --- | --- | --- | --- |
| Age (y) | 43.6 ± 7.2 | 44.4 ± 7.0 | 0.68 |
| BMI (kg/m<sup>2</sup>) | 23.5 ± 3.8 | 24.5 ± 3.2 | 0.32 |
| Body surface area (m<sup>2</sup>) | 1.68 ± 0.15 | 1.92 ± 0.16 | < 0.001 |
| Heart rate (beats/min) | 66.8 ± 9.5 | 63.4 ± 12.6 | 0.29 |
| LV parameters |  |  |  |
| End-diastolic volume (mL) | 118.4 ± 18.4 | 149.3 ± 23.5 | < 0.001 |
| End-diastolic volume index (mL/m<sup>2</sup>) | 70.5 ± 9.2 | 77.8 ± 10.5 | 0.01 |
| End-systolic volume (mL) | 40.3 ± 10.4 | 56.8 ± 14.3 | < 0.001 |
| End-systolic volume index (mL/m<sup>2</sup>) | 24.0 ± 5.8 | 29.6 ± 6.7 | 0.003 |
| Ejection fraction (%) | 66.0 ± 4.8 | 62.0 ± 5.3 | 0.007 |
| Stroke volume (mL) | 78.1 ± 11.6 | 92.5 ± 13.2 | < 0.001 |
| Mass (g) | 96.8 ± 15.3 | 138.2 ± 20.5 | < 0.001 |
| Mass index (g/m<sup>2</sup>) | 57.6 ± 7.4 | 72.0 ± 8.9 | < 0.001 |
| RV parameters |  |  |  |
| End-diastolic volume (mL) | 121.5 ± 21.3 | 154.8 ± 24.6 | < 0.001 |
| End-diastolic volume index (mL/m<sup>2</sup>) | 72.3 ± 10.1 | 80.6 ± 11.2 | 0.01 |
| End-systolic volume (mL) | 45.6 ± 12.5 | 62.8 ± 15.4 | < 0.001 |
| End-systolic volume index (mL/m<sup>2</sup>) | 27.1 ± 6.7 | 32.7 ± 7.5 | 0.01 |
| Ejection fraction (%) | 62.5 ± 5.3 | 59.4 ± 5.7 | 0.046 |
| Stroke volume (mL) | 75.9 ± 12.4 | 92.0 ± 13.8 | < 0.001 |
| Mass (g) | 24.8 ± 5.2 | 34.3 ± 6.1 | < 0.001 |
| Mass index (g/m<sup>2</sup>) | 14.8 ± 2.8 | 17.9 ± 3.0 | < 0.001 |
| Wall thickness parameters |  |  |  |
| Interventricular septal thickness (mm) | 7.9 ± 1.2 | 9.6 ± 1.3 | < 0.001 |
| LV posterior wall thickness (mm) | 7.4 ± 1.1 | 9.1 ± 1.2 | < 0.001 |

Note.—Unless otherwise specified, data are presented as means ± SDs. Healthy volunteer status was verified through 3.0-T bSSFP Cine scanning. BMI = body mass index, LV = left ventricle, RV = right ventricle, SD = standard deviation. *P* < 0.05 indicates statistical significance.

### Myocardial T1 and T2 Estimations

Fig. 2 displays representative T1 and T2 maps acquired using different sequences at 3.0 T and 5.0 T, where overall good mapping quality can be observed for all acquisitions without obvious artifacts. Compared to the bSSFP readout, the FLASH based mapping techniques at 3.0 T exhibits larger variations in the T1 and T2 maps, which is caused by the reduced SNR associated with FLASH acquisition. Comparing across the field strengths, T1 and T2 maps obtained with 5T-FLASH techniques are visually more smooth than the counterpart 3T-FLASH, albeit with reduced myocardium-to-blood contrast. For both field strengths, the T1 mapping quality of Multimap is slightly inferior to MOLLI. Table 3 summarizes myocardial T1 and T2 values obtained by different mapping sequences across field strengths and readouts, which are further compared in Fig. 3. At 3.0 T, MOLLI with FLASH readout yielded significantly higher T1 values (1305.9 ± 38.3 ms) compared to bSSFP readout (1159.9 ± 27.8 ms). With the field strength increasing to 5.0 T, the MOLLI FLASH T1 further increased to 1452.9 ± 33.2 ms. Under the same setting of field strength and readout, T1 values measured by Multimap were consistently and significantly higher than those measured by MOLLI. All inter-group comparisons for T1 measurements reached statistical significance (Fig. 3B, all *P* < 0.05).

**Fig. 2:**
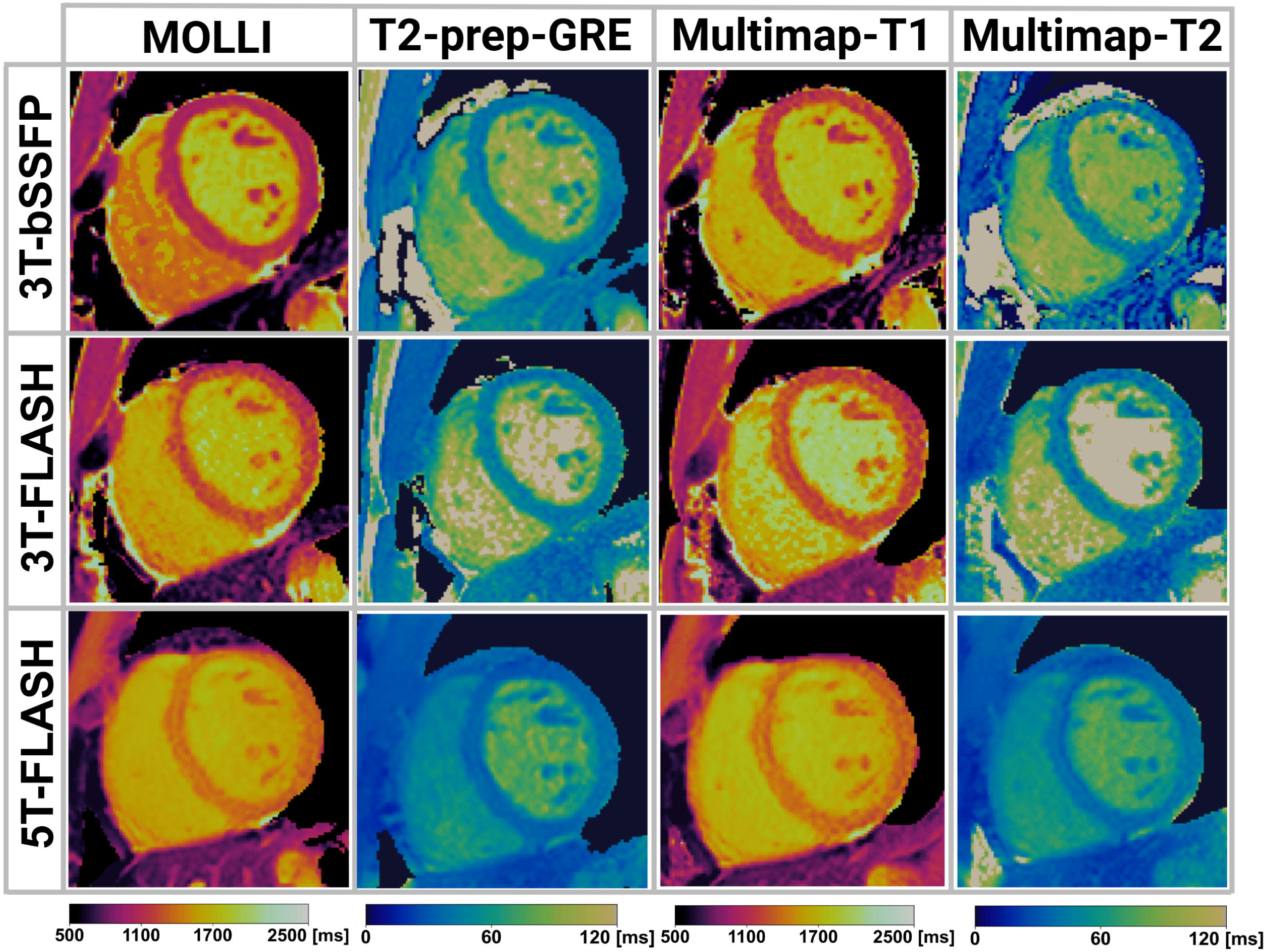
Representative T1 and T2 maps at 3.0-T and 5.0-T MRI using different mapping techniques. Both bSSFP and FLASH readouts were employed for MOLLI T1 mapping, T2-prep-GRE T2 mapping, and Multimap simultaneous T1 and T2 mapping (top and middle rows), while only the FLASH readout was adopted for 5.0-T mapping (bottom row). The mapping variations were reduced in 5T-FLASH compared with counterpart 3T-FLASH, albeit with reduced myocardium to blood contrast. MOLLI = MOdified Look Locker Inversion recovery, bSSFP = balanced steady-state free precession, Multimap = multi-parametric mapping, T2-prep-GRE = T2-prepared Gradient Echo, FLASH = Fast low-angle shot.

**Fig. 3:**
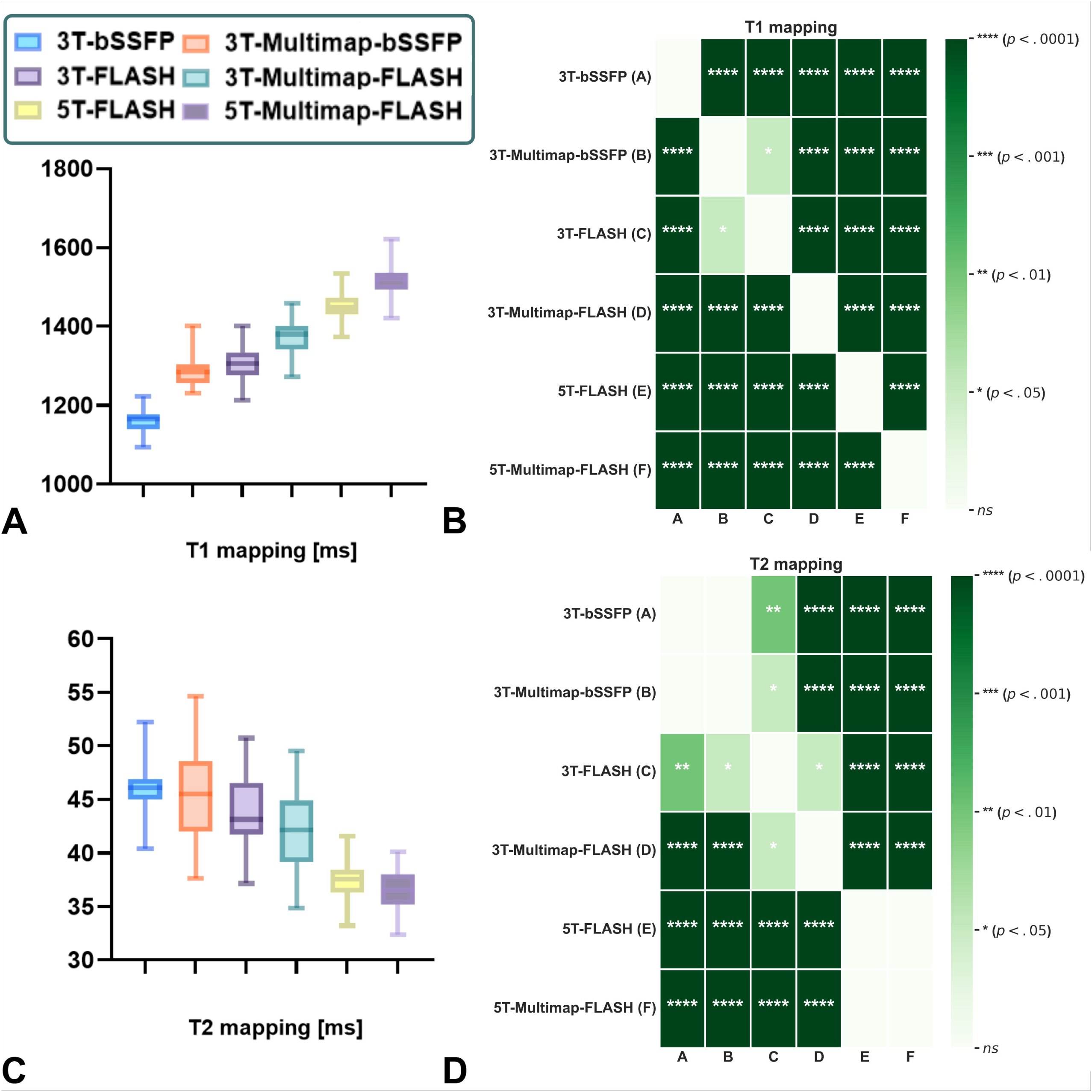
Comparison of myocardial T1 and T2 estimations across different field strengths and sequences. **(A, B)** Box plots show the distribution of native myocardial T1 and T2 values acquired with 3T-bSSFP, 3T-Multimap-bSSFP, 3T-FLASH, 3T-Multimap-FLASH, 5T-FLASH, and 5T-Multimap-FLASH. **(C, D)** Heatmaps indicate the statistical significance of pairwise comparisons among groups for T1 and T2 values based on one-way ANOVA followed by post hoc tests. The number of asterisks indicates the level of significance: *\*\*\*\*P* < 0.0001, *\*\*\*P* < 0.001; *\*\*P* < 0.01, *\*P* < 0.05, ns = not significant. bSSFP = balanced steady-state free precession, FLASH = Fast low-angle shot, Multimap = multi-parametric mapping.

**Table 3:** Comparison of Myocardial T1 and T2 Values Measured at 3.0-T and 5.0-T MRI.

| Field Strength (Readout) | MOLLI | T2-prep-GRE | Multimap-T1 | Multimap-T2 |
| --- | --- | --- | --- | --- |
| 3.0T (bSSFP) | 1159.9 ± 27.8 | 1283.8 ± 35.3 | 46.20 ± 1.97 | 45.64 ± 3.95 |
| 3.0T (FLASH) | 1305.9 ± 38.3 | 1373.7 ± 42.1 | 43.97 ± 2.95 | 42.10 ± 3.29 |
| 5.0T (FLASH) | 1452.9 ± 33.2 | 1515.2 ± 39.2 | 37.53 ± 1.74 | 36.72 ± 1.99 |

Note. — Unless otherwise specified, data are presented as means ± SDs. MOLLI = MOfified Look Locker Inversion recovery, T2-prep-GRE = T2-prepared Gradient Echo, bSSFP = balanced steady-state free precession, FLASH = Fast low-angle shot, Multimap = multi-parametric mapping, SD = standard deviation.

The magnetic field strength and readout type also have significant effect on T2 measurements (Fig. 3C, D). At 3.0 T, T2 values obtained with FLASH readout (43.97 ± 2.95 ms) were significantly lower than those with bSSFP readout (46.20 ± 1.97 ms) (*P* < 0.01). With the field strength increasing to 5.0 T, the measured T2 values further decreased to 37.53 ± 1.74 ms, significantly lower than 3T-FLASH (*P* < 0.0001). Multimap T2 measurements were consistently lower than T2-prep-GRE with the same readout for both field strengths. The heatmap matrix in Fig. 3D further reveals the pattern of statistical differences in T2 values between different sequences.

### Reproducibility Assessment

The CoV and ICC values assessing intra-observer, inter-observer, and scan-rescan reproducibility are summarized in Table 4. The intra- and inter-observer ICC was respectively greater than 0.89 and 0.82, indicating reliable T1 and T2 measurements. The readout affected measurement precision, with 3T-bSSFP demonstrating lower scan-rescan CoV and higher ICC than 3T-FLASH. The scan-rescan reproducibility analysis revealed that the CoV of 5T-FLASH (2.41% - 2.96%) was superior to 3T-FLASH (2.89% - 3.45%), and approached those of 3T-bSSFP (2.37% - 2.92%). The separate acquisition sequences (MOLLI and T2-prep-GRE) demonstrated better reproducibility than the counterpart Multimap with the same readout and field strength settings.

**Table 4:** Reproducibility Assessment of Cardiac Mapping at 3.0-T and 5.0-T MRI.

| Parameter | MOLLI<br>(bSSFP) | Multimap-T1 (bSSFP) | MOLLI<br>(FLASH) | Multimap-T1<br>(FLASH) | T2-prep-G<br>RE<br>(bSSFP) | Multimap-T2 (bSSFP) | T2-prep-G<br>RE<br>(FLASH) | Multimap-T2<br>(FLASH) |
| --- | --- | --- | --- | --- | --- | --- | --- | --- |
| 3.0-T Intra-observer |  |  |  |  |  |  |  |  |
| CoV (%; 95% CI) | 0.84<br>(0.70-1.00) | 0.98<br>(0.83-1.16) | 1.21<br>(1.03-1.43) | 1.35<br>(1.13-1.61) | 1.13<br>(0.95-1.33) | 1.31<br>(1.11-1.54) | 1.54<br>(1.29-1.84) | 1.67<br>(1.37-2.01) |
| ICC (95% CI) | 0.97<br>(0.95-0.98) | 0.95<br>(0.92-0.97) | 0.93<br>(0.90-0.95) | 0.92<br>(0.89-0.94) | 0.95<br>(0.93-0.97) | 0.92<br>(0.90-0.95) | 0.90<br>(0.88-0.93) | 0.89<br>(0.87-0.92) |
| 3.0-T Inter-observer |  |  |  |  |  |  |  |  |
| CoV (%; 95% CI) | 0.95<br>(0.78-1.17) | 1.13<br>(0.92-1.37) | 1.38<br>(1.14-1.66) | 1.53<br>(1.28-1.84) | 1.29<br>(1.07-1.56) | 1.49<br>(1.26-1.77) | 1.74<br>(1.46-2.08) | 1.89<br>(1.56-2.28) |
| ICC (95% CI) | 0.95<br>(0.92-0.97) | 0.92<br>(0.89-0.94) | 0.89<br>(0.86-0.92) | 0.87<br>(0.84-0.90) | 0.92<br>(0.89-0.94) | 0.89<br>(0.86-0.92) | 0.85<br>(0.81-0.89) | 0.82<br>(0.78-0.86) |
| 5.0-T Intra-observer |  |  |  |  |  |  |  |  |
| CoV (%; 95% CI) | - | - | 0.95<br>(0.81-1.13) | 1.12<br>(0.95-1.32) | - | - | 1.24<br>(1.05-1.46) | 1.43<br>(1.20-1.70) |
| ICC (95% CI) | - | - | 0.96<br>(0.94-0.97) | 0.94<br>(0.91-0.96) | - | - | 0.94<br>(0.92-0.96) | 0.91<br>(0.89-0.94) |
| 5.0-T Inter-observer |  |  |  |  |  |  |  |  |
| CoV (%; 95% CI) | - | - | 1.09<br>(0.89-1.32) | 1.28<br>(1.06-1.54) | - | - | 1.41<br>(1.17-1.70) | 1.61<br>(1.35-1.92) |
| ICC (95% CI) | - | - | 0.93<br>(0.90-0.95) | 0.90<br>(0.87-0.93) | - | - | 0.91<br>(0.88-0.93) | 0.87<br>(0.84-0.90) |
| 3.0T Scan-rescan |  |  |  |  |  |  |  |  |
| CoV (%; 95% CI) | 2.37<br>(1.97-2.83) | 2.64<br>(2.21-3.15) | 2.89<br>(2.41-3.46) | 3.13<br>(2.61-3.75) | 2.92<br>(2.44-3.49) | 3.21<br>(2.69-3.83) | 3.45<br>(2.88-4.12) | 3.72<br>(3.11-4.45) |
| ICC (95% CI) | 0.94<br>(0.91-0.96) | 0.92<br>(0.89-0.94) | 0.90<br>(0.87-0.93) | 0.88<br>(0.85-0.91) | 0.91<br>(0.88-0.93) | 0.89<br>(0.86-0.92) | 0.87<br>(0.84-0.90) | 0.85<br>(0.82-0.88) |
| 5.0T Scan-rescan |  |  |  |  |  |  |  |  |
| CoV (%; 95% CI) | - | - | 2.41<br>(2.01-2.88) | 2.68<br>(2.24-3.20) | - | - | 2.96<br>(2.47-3.54) | 3.25<br>(2.72-3.88) |
| ICC (95% CI) | - | - | 0.92<br>(0.89-0.94) | 0.90<br>(0.87-0.93) | - | - | 0.89<br>(0.86-0.92) | 0.87<br>(0.84-0.90) |

Note.—Unless otherwise indicated, data are presented as CoV (%) and ICC, with 95% CIs in parentheses. CoV = coefficient of variation, ICC = intraclass correlation coefficient, CI = confidence interval, MOLLI = MOdified Look Locker Inversion recovery, T2-prep-GRE = T2-prepared Gradient Echo, bSSFP = balanced steady-state free precession, FLASH = Fast low-angle shot, Multimap = multi-parametric mapping. - indicates not applicable.

### Agreement between 3.0-T and 5.0-T MR for Myocardial Mapping

The Bland-Altman analyses evaluating the agreement between 3.0 T and 5.0 T myocardial T1 and T2 mapping, and between Multimap and conventional separate mapping techniques at 5.0-T MRI are presented in Fig. 4. Myocardial T1 values measured at 5.0 T were higher than those measured at 3.0 T for both MOLLI (bias, 147.0 ms; 95% LOA: 50.9, 243.0 ms) and Multimap (bias, 141.5 ms; 95% LOA: 22.9, 260.1 ms). Conversely, myocardial T2 values obtained at 5.0 T were lower than at 3.0 T (bias, −6.4 ms; 95% LOA: −12.8, −0.1 ms for T2-prep-GRE; bias, −5.4 ms; 95% LOA: −13.2, 2.4 ms for Multimap). Regarding the different mapping techniques at 5.0 T, T1 values measured by the Multimap were higher than those from the MOLLI (bias, 62.3 ms; 95% LOA: −44.1, 168.7 ms), whereas the difference in T2 values between Multimap and T2-prep-GRE was minimal (bias, −0.8 ms; 95% LOA: −5.8, 4.2 ms). The Bland-Altman plots showed random distribution of data points around the mean difference line, with no apparent systematic trends.

**Fig. 4:**
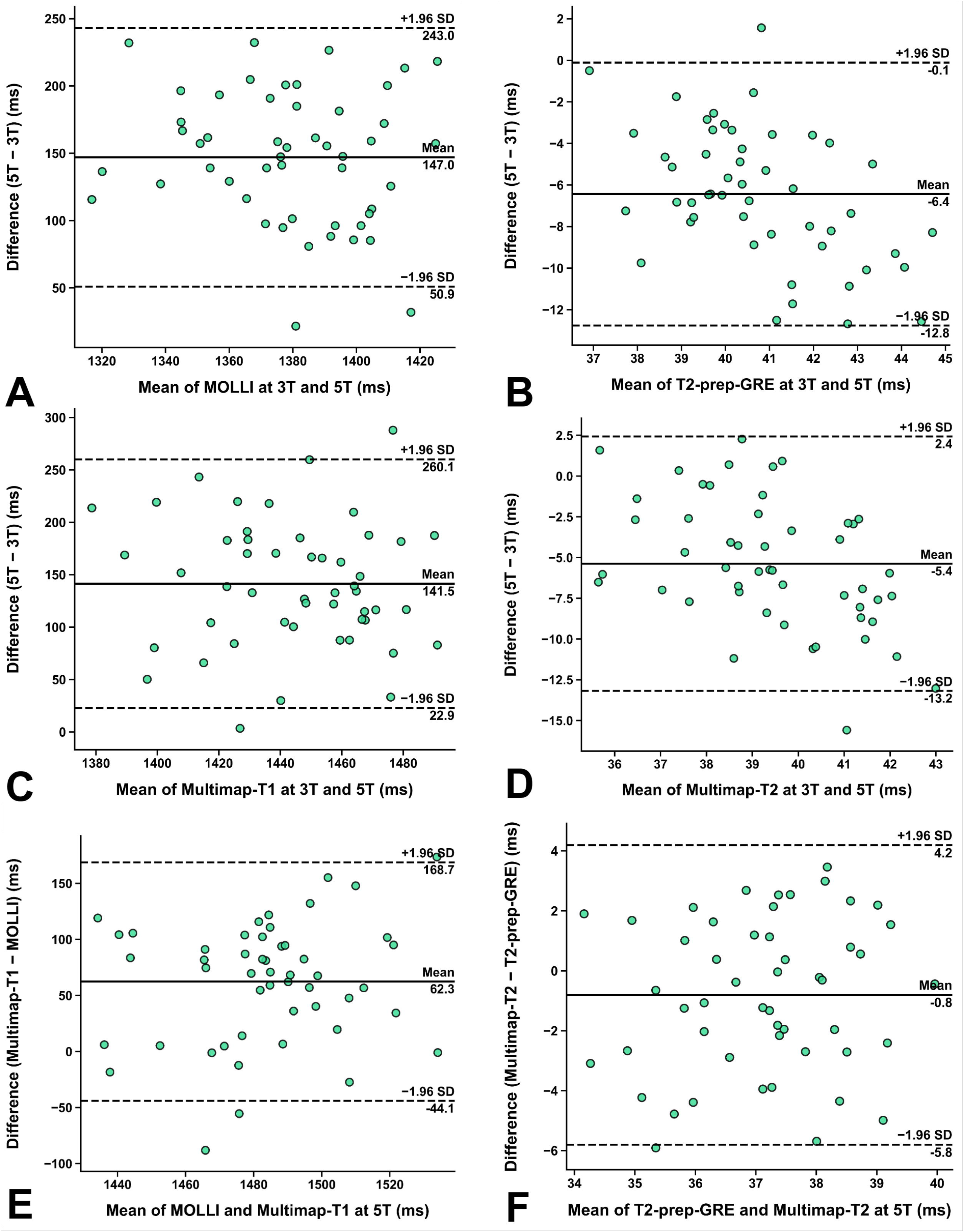
Scatterplots show the comparison of myocardial T1 and T2 values with Bland-Altman analysis. Scatterplots show **(A, B)** differences in T1 and T2 values between 5.0-T and 3.0-T mapping using MOLLI and T2-prep-GRE with the FLASH readout, **(C, D)** differences in T1 and T2 values between 5.0-T and 3.0-T MRI using the Multimap sequence with the FLASH readout, and **(E, F)** differences in T1 and T2 values between Multimap and MOLLI, T2-prep-GRE at 5.0-T. Center line represents mean of differences, top line shows upper 95% limit of agreement, and bottom line shows lower 95% limit of agreement. MOLLI = MOdified Look Locker Inversion recovery, T2-prep-GRE = T2-prepared Gradient Echo, Multimap = multi-parametric mapping, FLASH = Fast low-angle shot, SD = standard deviation.

### Segmental Analysis

Fig. 5 and Fig. 6 respectively illustrates the bullseye plots of the mean and SD of T1 and T2 measurements at 3.0 T and 5.0 T, where the number in each segment is obtained by averaging across all subjects.

**Fig. 5:**
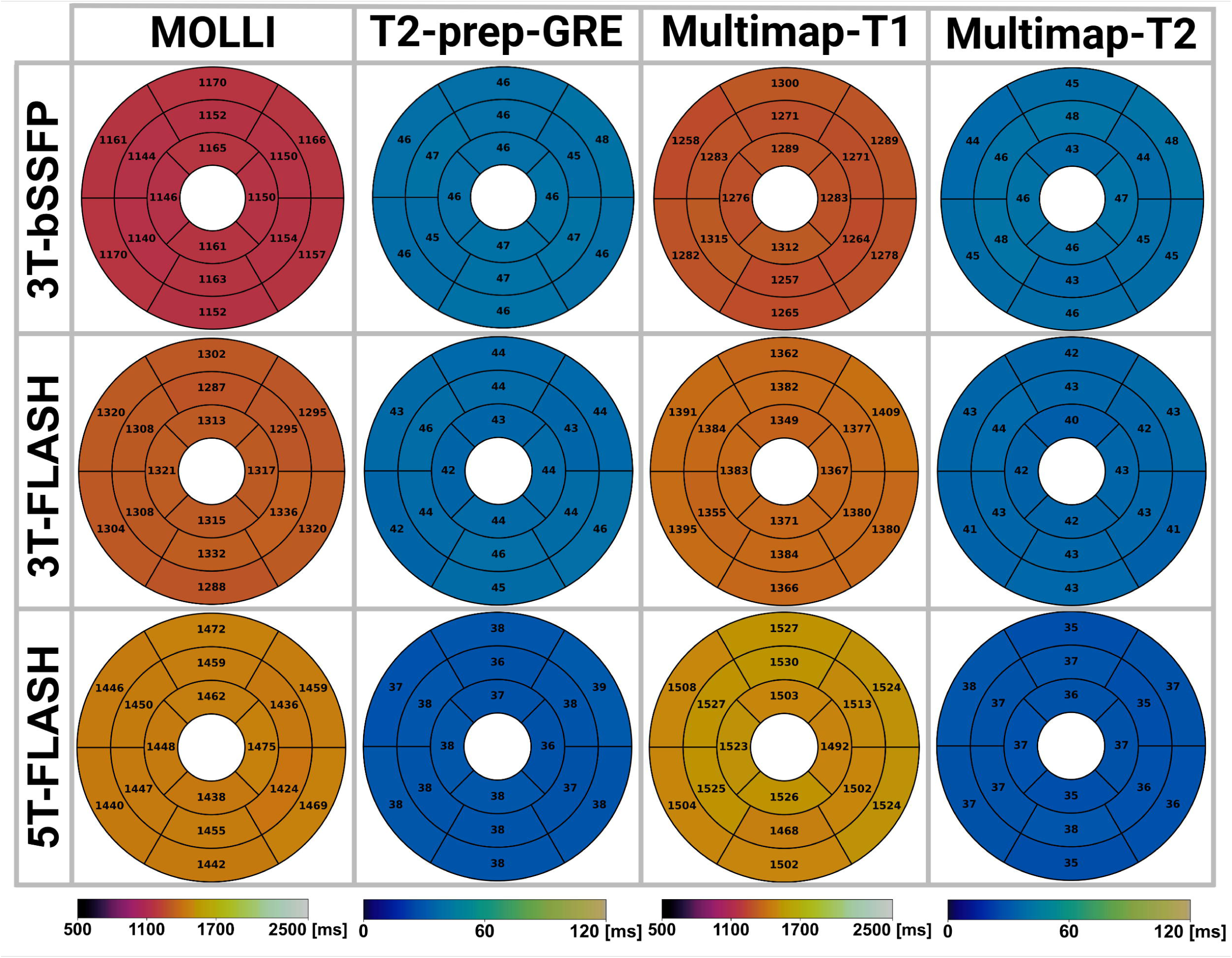
Bullseye plots of myocardial T1 and T2 measured at 3.0-T and 5.0-T MRI with different techniques, including separate mapping techniques of MOLLI and T2-prep-GRE, and simultaneous T1 and T2 mapping technique of Multimap. Both bSSFP and FLASH readouts were adopted for 3.0-T mapping, while only FLASH readout was employed for 5.0-T imaging. The value in each segment was obtained by averaging across all subjects. MOLLI = MOdified Look Locker Inversion recovery, bSSFP = balanced steady-state free precession, Multimap = multi-parametric mapping, T2-prep-GRE = T2-prepared Gradient Echo, FLASH = Fast low-angle shot.

**Fig. 6:**
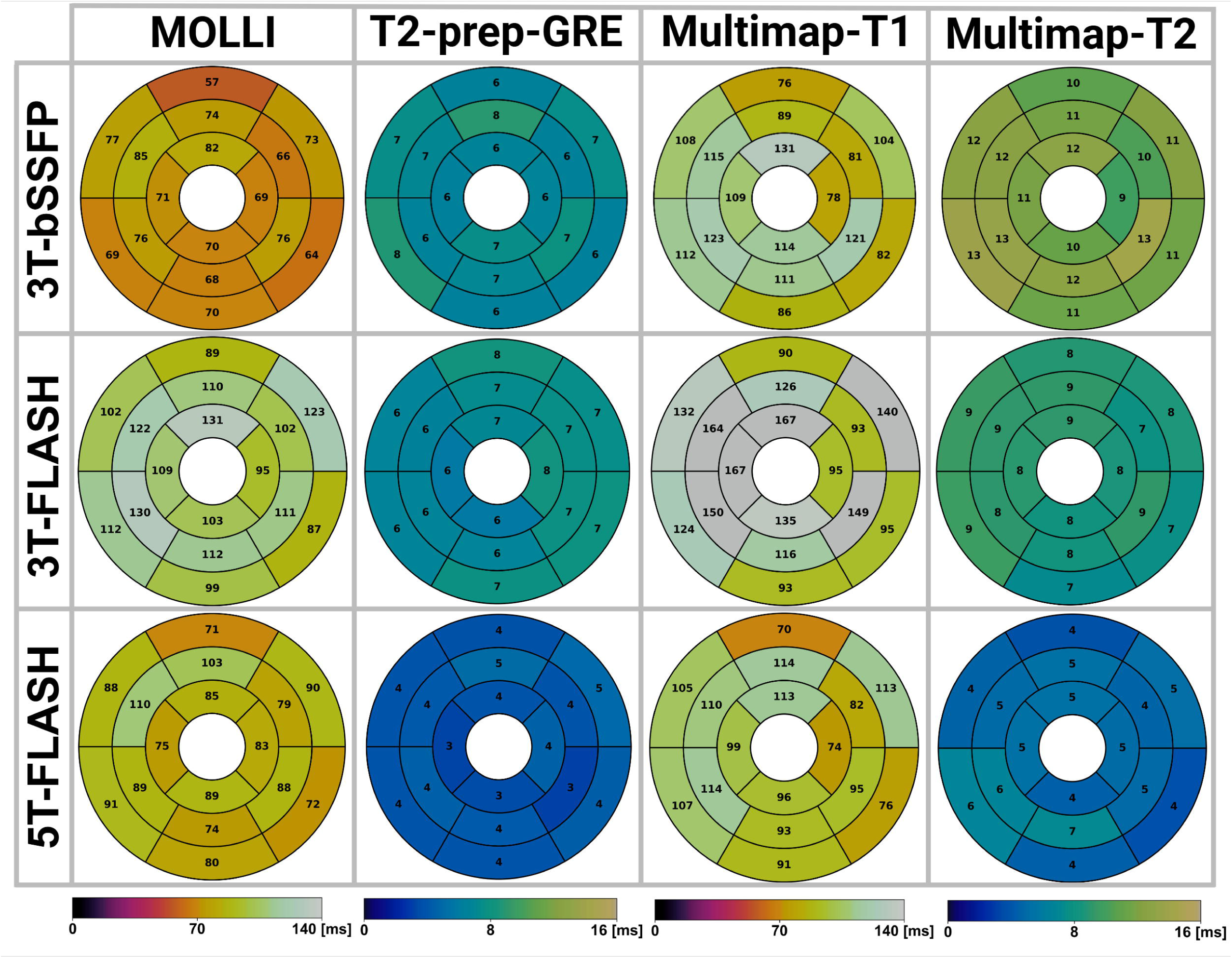
Bullseye plots of standard deviation of myocardial T1 and T2 measured at 3.0-T and 5.0-T MRI with different techniques, including separate mapping techniques of MOLLI and T2-prep-GRE, and simultaneous T1 and T2 mapping technique of Multimap. Both bSSFP and FLASH readouts were adopted for 3.0-T mapping, while only FLASH readout was employed for 5.0-T imaging. The value in each segment was obtained by averaging across all subjects. MOLLI = MOdified Look Locker Inversion recovery, bSSFP = balanced steady-state free precession, Multimap = multi-parametric mapping, T2-prep-GRE = T2-prepared Gradient Echo, FLASH = Fast low-angle shot.

The mean T1 distribution was overall homogeneous for MOLLI-based techniques at both field strengths, while Multimap-T1 exhibited slightly increased intersegmental variations compared to MOLLI, and these differences were similar between bSSFP and FLASH readouts. Regarding T1 measurement precision, 3T-bSSFP MOLLI and Multimap had lower intrasegment SDs than the corresponding 3T-FLASH. Multimap-T1 exhibited higher intrasegment SDs than MOLLI across all field strengths and readouts, indicating relatively lower measurement precision. Comparing across field strengths, the T1 SD of 5T-FLASH was lower than that of 3T-FLASH, but higher than that of 3T-bSSFP.

The mean myocardial T2 values were homogeneously distributed across all segments for all mapping techniques. The T2 measurement SD was the lowest for 5T-FLASH, followed by 3T-bSSFP and 3T-FLASH. With the bSSFP readout, Multimap-T2 exhibited greater measurement variability than T2-prep-GRE, while the SDs were similar between Multimap-T2 and T2-prep-GRE with the FLASH readout at both field strengths.

## Discussion

Cardiac T1 and T2 mapping have been extensively investigated at 1.5-T and 3.0-T MR, the study of which is limited for the whole-body 5.0-T scanner. This study systematically compared the performance of 3.0-T and 5.0-T MR for myocardial mapping using MOLLI, T2-prep-GRE, and Multimap. All mapping techniques at 5.0 T produced high-quality, artifact-free quantitative maps comparable to 3.0-T CMR mapping. Notably, the T1 and T2 measurement reproducibility and precision with 5T-FLASH were higher than 3T-FLASH, and were only slightly inferior to the 3.0-T mapping techniques with standard bSSFP readout. This study demonstrated the feasibility of conventional separate mapping techniques and Multimap at 5.0 T, and established reference T1 and T2 values for normal myocardium.

The 5.0-T whole-body scanner holds the promise for high SNR and/or spatial resolution imaging. However, the ultrahigh field strength also pose special challenges for CMR due to increased field inhomogeneities and radiofrequency power deposition. Recent studies have demonstrated the feasibility of late gadolinium enhancement and coronary CMR at 5.0 T, where 5.0-T MRI achieved image quality comparable or superior to 3.0-T MRI [14,18,22]. Compared to qualitative imaging, quantitative CMR is more vulnerable to hardware imperfections which warrants further investigation and optimization. To ensure effective and uniform inversion and T2 preparation in the parametric mapping acquisitions, the inversion pulse and the refocusing pulse in the T2-prep module have been optimized using numerical simulations and in phantoms, which was not in included in the current study due to space limitations. The finally adopted pulse for both 3.0-T and 5.0-T CMR was a tangent/hyperbolic tangent adiabatic pulse as also recommended by Kellman et al. due to its robustness to B0/B1+ inhomogeneities [23]. Furthermore, the B1+ spin history was modelled during dictionary simulation to correct flip errors attributed to inhomogeneous transmit B1 and imperfect slice profile, which has been demonstrated to improve the parameter estimation accuracy of the Multimap technique [11].

Myocardial T1 mapping has been demonstrated feasible at 5.0-T MR in a recent study using the FLASH MOLLI technique [15], reporting native T1 of ∼1500 ms, which is comparable to the value measured in this study. However, systematic comparison of the 5.0-T measurement precision and reproducibility with 3.0-T CMR was not performed in the previous study. Moreover, to the authors’ knowledge, myocardial T2 mapping at 5.0-T MR has not been investigated. We have demonstrated myocardial T2 mapping is feasible with the T2-prep-GRE and Multimap techniques, and found that increasing the field strength from 3.0-T to 5.0-T MR results in an 14.6% decrease in T2 values (43.97 ± 2.95 ms vs. 37.53 ± 1.74 ms) as measured with FLASH T2-prep-GRE, which should be attributed to enhanced susceptibility effects and B0/B1 inhomogeneity at higher field strength [24,25].

The scan-rescan CoV and ICC for 5T-FLASH were comparable to those of 3T-bSSFP and significantly better than those of 3T-FLASH. The T1 and T2 measurement SD of 5T-FLASH were also lower than 3T-FLASH and similar to those estimated with bSSFP-based techniques at 3.0 T. These results suggest that the SNR gain facilitated by 5.0 T could compensate for the SNR loss by using the FLASH readout in the parametric mapping acquisitions, leading to T1 and T2 mapping with good measurement reproducibility and precision. It is noted that denoising was not performed for any mapping technique at 3.0-T or 5.0-T MRI to validate its intrinsic mapping property, although denoising on the contrast weighted images before parameter estimation may aid in reducing measurement variability and improving mapping reproducibility and precision.

Comparison of mapping techniques at 5.0 T revealed obvious differences in T1 measurements between Multimap and MOLLI, while T2 measurement with MulitMap was similar to T2-prep-GRE. Similar trends have been observed for these techniques at 3.0 T. Further segmental analysis confirmed good spatial homogeneity of the 5.0-T cardiac parametric mapping. It is noted that the basal inferior and inferolateral segments showed higher SDs across different sequences and field strengths. This may be attributed to partial volume effects and susceptibility artifacts in these regions [16]. Additionally, Multimap quantifying T1 and T2 simultaneously in a short scan of ten heartbeats is more efficient than the separate mapping techniques, and its measurement precision as measured by SD was only slightly lower than MOLLI and T2-prep-GRE at both field strengths. Given the good reproducibility of Multimap (ICC > 0.8), it has great potential for 5.0-T CMR applications.

## Limitations

This study has several limitations. Firstly, only healthy subjects were recruited in this study to establish the reference T1 and T2 values for normal myocardium, and further investigation of 5.0-T CMR mapping in cardiomyopathy patients are warranted. Secondly, contrast injection is not allowed in our center, and post-contrast T1 mapping was not performed in this study. Thirdly, this was a single-center investigation, which may be subject to center-specific and manufacturer-specific biases. The relatively limited sample size also suggests that the generalizability of this study’s findings requires validation through multi-center studies with larger cohorts.

## Conclusions

In conclusion, this study demonstrated the feasibility and good reproducibility of MOLLI, T2-prep-GRE, and Multimap at 5.0-T MRI, and established preliminary reference T1 and T2 values for normal myocardium. These findings are significant for the development of quantitative CMR at 5.0-T, serving as a foundation for establishing normal standards and understanding parameter distributions in clinical applications.

## Data Availability

All data produced in this study are available from the corresponding author upon reasonable request.

## Abbreviations

AHA: American Heart Association
bSSFP: balanced steady-state free precession
CMR: cardiovascular magnetic resonance
CoV: coefficient of variation
FLASH: Fast low-angle shot
GRAPPA: generalized autocalibrating partially parallel acquisition
ICC: intraclass correlation coefficient
LOA: limits of agreement
MOLLI: MOdified Look Locker Inversion recovery
SD: standard deviation
SNR: signal-to-noise ratio
Multimap: multi-parametric mapping
T2-prep-GRE: T2-prepared Gradient Echo

## Declarations

### Ethics approval and consent to participate

This study was approved by the institutional ethics committee of ShanghaiTech University. All participants provided written informed consent prior to enrollment. The study was conducted in accordance with the Declaration of Helsinki. No animal studies were conducted.

### Consent for publication

Not applicable.

### Availability of data and materials

The datasets used in this study are available from the corresponding author upon reasonable request.

### Authors’ contributions

**Haikun Qi:** Conceptualization, Funding acquisition, Methodology, Project administration, Supervision, Resources, Writing - review & editing. **Peng Hu:** Supervision, Writing - review & editing, Resources. **Tianyi Wu:** Data curation, Formal analysis. **Wenjian Liu:** Software, Investigation, Data curation, Validation. **Zhenfeng Lyu:** Methodology, Data curation, Investigation, Software. **Hongzhang Huang:** Data curation, Investigation, Methodology, Software, Validation, Visualization, Writing - review & editing. **Qinfang Miao:** Data curation, Formal analysis, Investigation, Methodology, Validation, Visualization, Writing - original draft, Writing - review & editing.

## Acknowledgments

Not applicable.

## References

[1] Dr M, Jc M, Vm F, L G-W, T H, P K, et al. Clinical recommendations for cardiovascular magnetic resonance mapping of T1, T2, T2* and extracellular volume: A consensus statement by the Society for Cardiovascular Magnetic Resonance (SCMR) endorsed by the European Association for Cardiovascular Imaging (EACVI). J Cardiovasc Magn Reson Off J Soc Cardiovasc Magn Reson 2017;19. 10.1186/s12968-017-0389-8.

[2] W W, A A-A, A S, P T, Rm W, M P, et al. Clinical Impact of Cardiac MRI T1 and T2 Parametric Mapping in Patients with Suspected Cardiomyopathy. Radiology 2022;305. 10.1148/radiol.220067.

[3] Taylor AJ, Salerno M, Dharmakumar R, Jerosch-Herold Michael. T1 Mapping. JACC Cardiovasc Imaging 2016;9:67–81. 10.1016/j.jcmg.2015.11.005.

[4] Puntmann VO, Peker E, Chandrashekhar Y, Nagel E. T1 Mapping in Characterizing Myocardial Disease. Circ Res 2016;119:277–99. 10.1161/CIRCRESAHA.116.307974.

[5] Nakamori S, Dohi K, Ishida M, Goto Y, Imanaka-Yoshida K, Omori T, et al. Native T1 Mapping and Extracellular Volume Mapping for the Assessment of Diffuse Myocardial Fibrosis in Dilated Cardiomyopathy. JACC Cardiovasc Imaging 2018;11:48–59. 10.1016/j.jcmg.2017.04.006.

[6] Puntmann VO, Carr-White G, Jabbour A, Yu C-Y, Gebker R, Kelle S, et al. T1-Mapping and Outcome in Nonischemic Cardiomyopathy: All-Cause Mortality and Heart Failure. JACC Cardiovasc Imaging 2016;9:40–50. 10.1016/j.jcmg.2015.12.001.

[7] O’Brien AT, Gil KE, Varghese J, Simonetti OP, Zareba KM. T2 mapping in myocardial disease: a comprehensive review. J Cardiovasc Magn Reson Off J Soc Cardiovasc Magn Reson 2022;24:33. 10.1186/s12968-022-00866-0.

[8] Cau R, Falconi G, Suri JS, Saba L. T2 Mapping in Acute Myocarditis: Advancing Quantitative CMR Imaging for Precision Medicine. Clin Radiol 2025:107007. 10.1016/j.crad.2025.107007.

[9] Messroghli DR, Radjenovic A, Kozerke S, Higgins DM, Sivananthan MU, Ridgway JP. Modified Look-Locker inversion recovery (MOLLI) for high-resolution T1 mapping of the heart. Magn Reson Med 2004;52:141–6. 10.1002/mrm.20110.

[10] Giri S, Chung Y-C, Merchant A, Mihai G, Rajagopalan S, Raman SV, et al. T2 quantification for improved detection of myocardial edema. J Cardiovasc Magn Reson 2009;11:56. 10.1186/1532-429X-11-56.

[11] Lyu Z, Hua S, Xu J, Shen Y, Guo R, Hu P, et al. Free-breathing simultaneous native myocardial T1, T2 and T1ρ mapping with Cartesian acquisition and dictionary matching. J Cardiovasc Magn Reson Off J Soc Cardiovasc Magn Reson 2023;25:63. 10.1186/s12968-023-00973-6.

[12] Henningsson M. Cartesian dictionary-based native T1 and T2 mapping of the myocardium. Magn Reson Med 2022;87:2347–62. 10.1002/mrm.29143.

[13] Miao Q, Hua S, Gong Y, Lyu Z, Qian P, Liu C, et al. Free-breathing non-contrast T1ρ dispersion magnetic resonance imaging of myocardial interstitial fibrosis in comparison with extracellular volume fraction. J Cardiovasc Magn Reson Off J Soc Cardiovasc Magn Reson 2024;26:101093. 10.1016/j.jocmr.2024.101093.

[14] Guo Y, Lin L, Zhao S, Sun G, Chen Y, Xue K, et al. Myocardial Fibrosis Assessment at 3-T versus 5-T Myocardial Late Gadolinium Enhancement MRI: Early Results. Radiology 2024;313:e233424. 10.1148/radiol.233424.

[15] Ge L, Zhang Y, Zhu H, Zhang L, Zhou Y, Wang H, et al. MyocardialT1mapping at 5T using multi-inversion recovery real-time spoiled GRE. Phys Med Biol 2025;70. 10.1088/1361-6560/add986.

[16] Guo Y, Lin L, Xu K, Zhao S, Sun G, Chen Y, et al. Myocardial native T1 and extracellular volume measurements at 5T: Feasibility study and initial experience. J Cardiovasc Magn Reson Off J Soc Cardiovasc Magn Reson 2025;27:101896. 10.1016/j.jocmr.2025.101896.

[17] Lin L, Liu P, Sun G, Wang J, Liang D, Zheng H, et al. Bi-ventricular assessment with cardiovascular magnetic resonance at 5 Tesla: A pilot study. Front Cardiovasc Med 2022;9:913707. 10.3389/fcvm.2022.913707.

[18] Qian X, Wang S, Wu Y, Miao X, Chen Y, Lu H, et al. Late Gadolinium Enhancement of Nonischemic Cardiomyopathy at 5.0 T versus 3.0 T: A Crossover Design Study. Radiol Cardiothorac Imaging 2024;6:e240035. 10.1148/ryct.240035.

[19] Heinrich MP, Jenkinson M, Bhushan M, Matin T, Gleeson FV, Brady SM, et al. MIND: modality independent neighbourhood descriptor for multi-modal deformable registration. Med Image Anal 2012;16:1423–35. 10.1016/j.media.2012.05.008.

[20] Bustin A, Lima da Cruz G, Jaubert O, Lopez K, Botnar RM, Prieto C. High-dimensionality undersampled patch-based reconstruction (HD-PROST) for accelerated multi-contrast MRI. Magn Reson Med 2019;81:3705–19. 10.1002/mrm.27694.

[21] Cerqueira MD, Weissman NJ, Dilsizian V, Jacobs AK, Kaul S, Laskey WK, et al. Standardized myocardial segmentation and nomenclature for tomographic imaging of the heart. A statement for healthcare professionals from the Cardiac Imaging Committee of the Council on Clinical Cardiology of the American Heart Association. Circulation 2002;105:539–42. 10.1161/hc0402.102975.

[22] Lu H, Miao X, Wang D, Zheng X, Zhang S, Wang R, et al. Feasibility and Clinical Application of 5-T Noncontrast Dixon Whole-Heart Coronary MR Angiography: A Prospective Study. Radiology 2024;313:e240389. 10.1148/radiol.240389.

[23] Kellman P, Herzka DA, Hansen MS. Adiabatic inversion pulses for myocardial T1 mapping. Magn Reson Med 2014;71:1428–34. 10.1002/mrm.24793.

[24] Akçakaya M, Basha TA, Weingärtner S, Roujol S, Berg S, Nezafat R. Improved quantitative myocardial T2 mapping: Impact of the fitting model. Magn Reson Med 2015;74:93–105. 10.1002/mrm.25377.

[25] Topriceanu C-C, Pierce I, Moon JC, Captur G. T2 and T2⁎ mapping and weighted imaging in cardiac MRI. Magn Reson Imaging 2022;93:15–32. 10.1016/j.mri.2022.07.012.

